# COVID-19 onset, stay-at-home orders, and racialized inequities in homicide mortality across the US

**DOI:** 10.64898/2026.04.28.26351994

**Authors:** Maryam Tanveer, N. Jeanie Santaularia Gomez, Kate Vinita Fitch, George M. Holmes, Kathryn E. Moracco, Michael D. Fliss, Naoko Fulcher, Shabbar I. Ranapurwala

## Abstract

We examined the impact of COVID-19 pandemic onset (2020 April) on homicide mortality in the United States.

We conducted a single interrupted time series analysis using homicide events from the National Vital Statistics System that occurred over six years (2017-2022), with COVID-19 onset as an interruption. Monthly homicide deaths rates were calculated per 100,000 person-years to create a monthly time series. We used autoregressive integrated moving average regression, adjusted for seasonality, to model the immediate and sustained trend changes in the homicide mortality rate ratios due to the pandemic. We stratified models by length of stay-at-home order, race and ethnicity, sex, age, and weapon used to examine effect measure modification.

In Jan 2017, the US homicide mortality rate was 5.9/100,000 PY. While there were annual seasonal changes, the overall time trend before April 2020 was stable. However, with COVID-19 onset, the overall homicide mortality rate ratio increased by 32% (95% CI: 0.23, 0.41), which persisted through 2022 without additional trend changes, but with seasonal variations. Immediate increases with stable sustained trends in homicide rates were also observed in most stratified analyses.

COVID-19 pandemic onset is associated with US homicide mortality rates immediately increasing and remaining stable and higher afterwards.

## Introduction

To contain the swiftly spreading, highly contagious COVID-19 respiratory virus in the United States (US), a national emergency was issued on March 13, 2020, which led to many states issuing stay-at-home orders (SAHOs) to slow transmission.^1^ It is hypothesized that the national emergency and SAHOs led to deep uncertainty, isolation, and social unrest as people experienced job loss, grief, and restrictions on travel and socialization.^2^ These stressors contributed to the substantial rise in substance abuse and overdose,^3^ economic and housing insecurity,^4^ and negative mental health outcomes,^5^ which in turn may have served a pathway to the initial rise in community violence following pandemic onset,^6,7^ with differential impacts among minoritized people. The rise in firearm sales from March 2020 through 2021, also driven by the social and political unrest due to the pandemic, may also have been a contributing factor to the increase in community violence following the COVID-19 pandemic onset.^8^ However, questions remain about the long-term impact of the COVID-19 pandemic and SAHOs on community violence.

Early studies found that the COVID-19 pandemic onset was associated with heightened racial, ethnic, and gender inequities in violence-related harms. In Connecticut there was a 55% increase in community violence after the COVID-19 pandemic started, with racial and ethnic minority groups experiencing a 61% greater increase compared to white communities.^9^ Another study found that the firearm homicide rate ratio increased between Black and White people with Black individuals experiencing a firearm homicide rate 11 times higher than White individuals in 2018 compared to 15 times higher in 2021.^10^ Comparing Black males aged 10-24 years to White males of the same age, the firearm homicide rate ratio increased from 20.6 in 2019 to 21.6 in 2020, with the homicide death rate for White males aged 10-24 being 0.9 homicide deaths per 100,000 persons.^11^ Among children, there was an increase in fatal and non-fatal firearm injury and abuse-related hospital treatments.^12^ And, among pregnant women, homicide rates increased by 41.2% from 2019 to 2020.^13^

Most reports on the impacts of the pandemic on community violence focused on 2020 alone and on specific states or specific outcomes rather than homicide mortality. It is unclear how the increased presence of firearms in communities because of increased sales during the pandemic impacted long-term post-COVID community violence trends in the US. Further, it is unknown how the different SAHO durations may have changed the impact of COVID-19 on community violence outcomes, specifically homicide mortality, and if the early inequities persisted beyond the first year of the pandemic.

To address these questions, we examined the long-term impact of COVID-19 pandemic following onset (April 2020) on homicide mortality in all 50 states of the US using data from January 1st, 2017 through December 31st, 2022, and examined effect measure modification by SAHOs, race and ethnicity, age, and sex.

## Methods

We conducted a quasi-experimental single interrupted time series analysis to examine the change in the trends of homicide mortality trends on the absolute and relative scale in all 50 states using data from the National Vital Statistics System (NVSS) from January 1st, 2017, through December 31st, 2022. We used deidentified individual death files from the NVSS that included dates of death, age at death, sex, race and ethnicity, the state in which death occurred, and international classification of disease (ICD) codes, 10^th^ revision each deceased individual. We also used Ballotpedia data^14^ to aggregate state-level information on SAHOs.

### Outcome Data

We identified homicide mortality using ICD 10 codes X85-Y09, Y87.1. We subdivided the homicide mortality outcome into firearm (ICD10: X93-95)^15^ and non-firearm (ICD10: X85-92, X96-Y09) homicide mortality. To calculate homicide mortality rate, we used annual state population estimates from 2017 through 2022 using NVSS bridged-race postcensal population data. However, the 2021 and 2022 population estimates were not available, hence, we used the 2020 population data to substitute the missing population estimates.

We estimated monthly homicide death rates per 100,000 person-years (PY) to create a monthly time series with 72 time points over six years for each of the 50 states. We also stratified our data to create time series of homicide mortality rates by age, sex, race and ethnicity, SAHOs, and firearm and non-firearm homicide mortality. We used monthly homicide death rates to examine absolute changes to homicide death rate trends.

### Exposure

The main exposure to this study was the COVID-19 pandemic. For the analysis on the absolute scale, the pre-pandemic period was between January 1st, 2017 through March 31st, 2020 (39 months). We considered April 1, 2020 (month 40) to be the onset of the COVID-19 pandemic onset, such that the pandemic period stretched from April 1st, 2020 through December 31st, 2022 (33 months).

For analysis on the relative scale, the pre-pandemic period was between July 1st, 2017 through March 31st, 2020 (33 months). This is because we used the average mortality rate from the first six months of 2017 as the baseline rate of homicide to compare changes following the onset of the COVID-19 pandemic. Again, for the relative scale analysis, we considered April 1st, 2020 (month 34) to be the onset of the COVID-19 pandemic onset, such that the pandemic period stretched from April 1st, 2020 through December 31st, 2022 (33 months).

### Covariates

The covariates used to examine effect modification of the impact of COVID-19 on homicide mortality rates were age (<18 years, 18-25, 26-35, 36-45, 46-55, 56-65, 66-75, 76 years or more), sex (male, female), race and ethnicity (Hispanic, non-Hispanic American native, non-Hispanic Asian, non-Hispanic black, and non-Hispanic white), SAHOs (states that had no SAHO, N=7; states that had a SAHO for less than a month, N=9; states that had a SAHO for more than a month, N=34; See supplementary materials for list of states), and use of firearm as the method of homicide (firearm and non-firearm).

### Statistical Analysis

We estimated overall and covariate stratified rate ratios (comparing monthly homicide rates beginning July 1, 2017 to the average homicide rate from January 1, 2017 through June 30, 2017) and homicide mortality rates during the pre-pandemic and pandemic periods along with 95% confidence intervals (CI) to examine baseline differences in firearm and non-firearm homicide mortality rates by covariates across the US.

We conducted single series interrupted time series (sITS) using autoregressive integrated moving average (ARIMA) regression to model the immediate and sustained trend changes in the United States homicide mortality rate ratios and rates, overall and by firearm use, after the start of the pandemic state of emergency, April 2020. Analysis was conducted using R programming language in RStudio. The astsa package was used to determine the parameters needed for the ARIMA model and seasonality.^16^ Seasonality was addressed with a harmonic term to account for the annual seasonality often observed in homicide data in the warmer summer months using the tsModel package.^17,18^

For the analysis on the absolute scale, the single series ITS model can be written as:

Homicide mortality rate/ rate ratio_COVID(p) x time(t)_= β0 + β1*time_t_ + β2*level_p_ + β3*time*level_p*t_ + β4*Harmonic-time (1,12) + e

Where, time is a continuous variable for the number of months in the time series (1-72 for mortality rate; 1-66 for rate ratio), level is a binary variable which is set to ‘0’ in the pre-pandemic period (Jan 2017-Feb 2020), and ‘1’ in the pandemic period (Mar 2020-Dec 2022). Further, β0, represents the homicide mortality rate or rate ratio in January 2017, β1 represents the time trends in homicide mortality rates or rate ratio pre-pandemic, β2 represents the immediate absolute change in the homicide mortality rate or rate ratio observed in April 2020 with the onset of the pandemic, β3 represent the change in trend of homicide mortality rates or rate ratio from pre-pandemic to the pandemic period and β4 represents a harmonic term to account for seasonality.

We stratified the models by race and ethnicity, sex, age, and SAHO to examine effect modification. Since the overall homicide rates varied considerably in some of the strata, to ensure comparison, we calculated homicide death rate ratios at each time point relative to the baseline homicide rate for the strata, where baseline homicide death rate is defined as the average of first six months (Jan 2017-Jun 2017) of homicide death rate. This resulted in another monthly time series with 66 time points over six years for each of the 50 states beginning in July 1, 2017 through December 31, 2022. The overall and stratified analyses were conducted on all homicide mortality trends and separately on firearm-related homicide and non-firearm-related homicide mortality. In the main text, we present stratified analysis on the relative scale, and results on the absolute scale are presented in the supplementary materials.

## Results

### Overall Analysis

During the study period (2017-2022), there were 137,610 homicide deaths in the United States. Before the pandemic, January 2017 to June 2017, the national average homicide mortality rate was 6.10 deaths per 100,000 PY. This mortality rate remained constant at around 6.10 deaths per 100,000 PY until April 2020 (Figure 1) and is used as the comparison reference value for the national rate ratio. The national average homicide death mortality rate increased in the period following the pandemic to 8.03 deaths/100,000 PY between April 2020 through December 2022.

**Figure 1.**
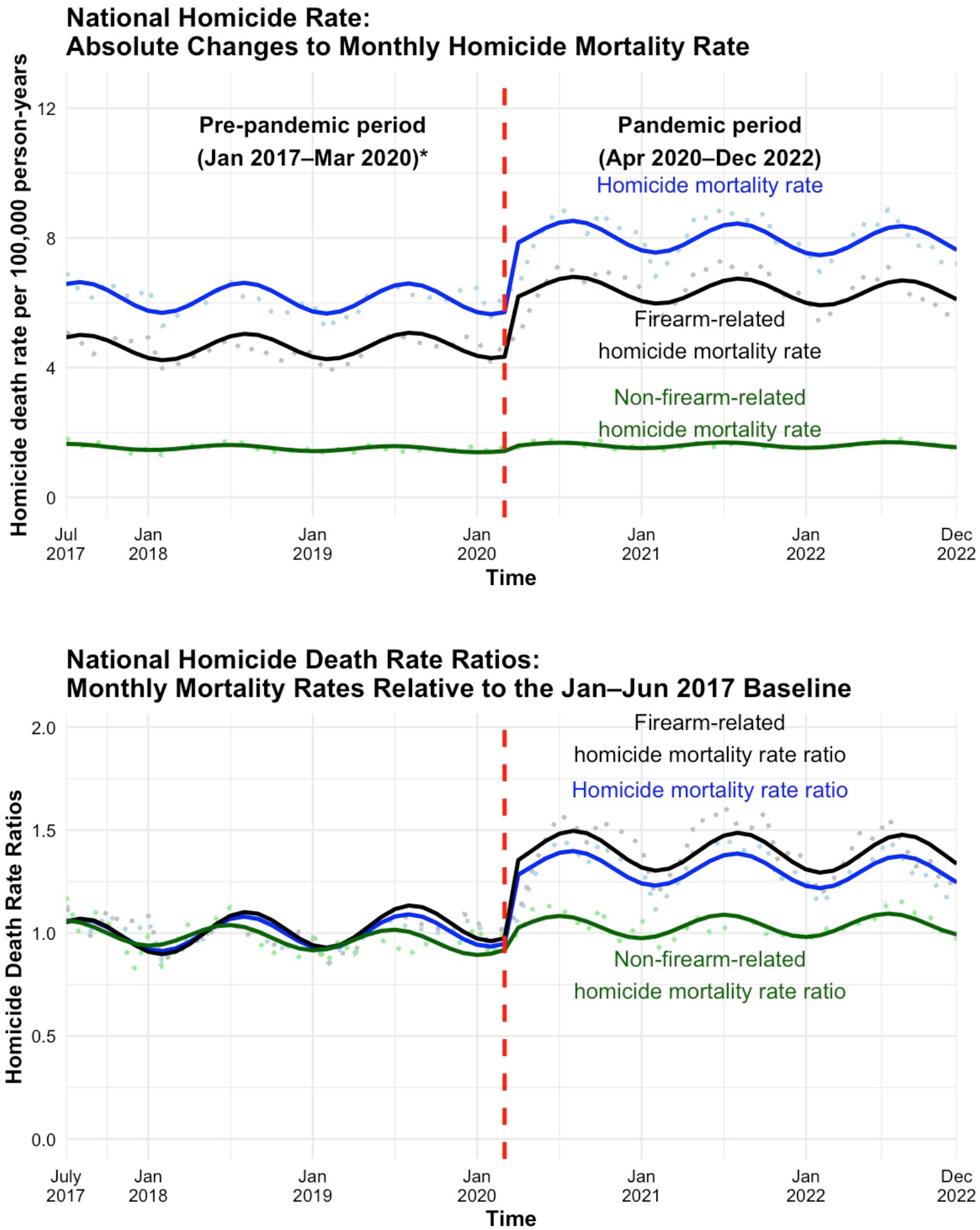
United States Homicide Death (a) Rates and (B) Rate Ratios Over Time per 100,000 PY, July 2017-December 2022. Legend: Dots represent raw monthly rates, and the lines represent the fitted monthly rate trends based on ARIMA modeling

Because the overall mortality rate from July 1st, 2017 through March 31st, 2020 was stable (Figure 1) and close to the reference homicide mortality rate (6.10 deaths per 100,000 PY), and since the pre-pandemic national average homicide mortality rate trend was stable (neither increasing or decreasing), the pre-pandemic overall rate ratio was 1.00. However, with COVID-19 pandemic onset in April 2020, the national homicide mortality rate immediately increased by 32% (95% CI: 23%, 41%) to 8.42 homicide deaths per 100,000 PY in May 2020 (Figure 1; Table 2). This trend persisted until the end of the study period with minimal trend changes. As a result, the average national homicide mortality rate ratio increased to 1.32 post-pandemic when comparing homicide mortality rates from April 1^st^, 2020 through December 31st, 2022 to the reference pre-pandemic mortality rate.

Similarly, immediate increases with the onset of the COVID-19 pandemic in April 2020 were observed for firearm-related homicide mortality rate ratios (35% (95% CI: 23%, 47%) and non-firearm-related homicide mortality rate ratios (8% (95%: 2%, 14%)), indicating that firearms were the primary method by which homicides increased. Both firearm- and non-firearm-related homicide mortality rate trends remained stable during the pandemic period (Figure 1; Table 2).

### Effect Modification Analysis

Stratifying by demographic factors, we compared each group’s post-pandemic onset homicide mortality rates to the same group’s 2017 pre-pandemic levels. We observe that groups with higher average rates of homicide pre-pandemic continue to face higher levels of average homicide mortality rates compared to their peers (Table 1). Non-Hispanic Black people experienced the highest rates of homicide death compared to all other racial and ethnic groups before (22.93 homicide deaths/100,000 PY) and post-pandemic (32.44 homicide deaths/100,000 PY). Similarly, higher homicide death rates were observed among males compared to females, and among 18 to 25-year-olds compared to other age groups pre- and post-pandemic (see Table 1). Additionally, states with SAHOs that lasted less than a month had the highest homicide mortality rates before (8.04 homicide deaths/100,000 PY) and after the pandemic began (10.67 homicide deaths/100,000 PY) compared to states with no SAHO (before pandemic: 4.13 homicide deaths/100,000 PY, after pandemic: 5.15 homicide deaths/100,000 PY) or SAHO that lasted a month or more (before pandemic: 5.41 homicide deaths/100,000 PY, after pandemic: 7.22 homicide deaths/100,000 PY).

**Table 1.** Average United States Homicide Rate per 100,000 PY Before and After COVID-19 Pandemic Initiation in March 2020, January 2017-December 2022.

|  | **Pre-COVID-19 Pandemic**<br>**Mean Homicide Rate per**<br>**100,000 PY**<br>**January 1st, 2017 - March**<br>**31st, 2020** | **Post-COVID-19 Pandemic**<br>**Mean Homicide Rate per**<br>**100,000 PY**<br>**April 1st, 2020 - December**<br>**31st, 2022\*** |
| --- | --- | --- |
| National Homicide | 6.10 | 8.03 |
| National Firearm | 4.58 | 6.42 |
| National Non-Firearm | 1.52 | 1.61 |
| Hispanic | 6.29 | 8.17 |
| Non-Hispanic American Native | 10.81 | 13.17 |
| Non-Hispanic Asian | 1.77 | 1.80 |
| Non-Hispanic Black | 22.89 | 32.44 |
| Non-Hispanic White | 2.78 | 3.11 |
| Female | 2.43 | 2.96 |
| Male | 9.88 | 13.26 |
| Under 18 | 2.27 | 3.15 |
| 18 to 25 | 13.96 | 18.40 |
| 26 to 35 | 11.86 | 15.99 |
| 36 to 45 | 8.18 | 11.38 |
| 46 to 55 | 5.17 | 6.69 |
| 56 to 65 | 3.57 | 4.30 |
| 66 to 75 | 2.18 | 2.51 |
| 76 or more | 1.88 | 2.03 |
| A month or more of stay-at-home order | 5.42 | 7.22 |
| Less than a month of stay-at-home order | 7.99 | 10.67 |
| Stay-at-home order not implemented | 4.11 | 5.15 |

**Table 2.** United States Homicide Risk Ratio Trend Changes Before and After COVID-19 Pandemic Initiation in March 2020, January 2017-December 2022.

|  | **Relative immediate change**<br>**when COVID-19 pandemic**<br>**started (April 1st, 2020)** |  | **Sustained trend change**<br>**during COVID-19**<br>**pandemic** |  |
| --- | --- | --- | --- | --- |
| National Homicide | 0.31 | [0.22, 0.39] | 0 | [-0.01, 0.00] |
| National Firearm | 0.35 | [0.23, 0.47] | 0 | [-0.01, 0.00] |
| National Non-Firearm | 0.08 | [0.02, 0.14] | 0 | [-0.00, 0.01] |
| Hispanic | 0.16 | [0.05, 0.26] | 0.01 | [0.00, 0.01] |
| Non-Hispanic American<br>Native | 0.13 | [-0.16, 0.42] | 0 | [-0.02, 0.01] |
| Non-Hispanic Asian | 0.03 | [-0.12, 0.18] | -0.01 | [-0.01, 0.00] |
| Non-Hispanic Black | 0.42 | [0.30, 0.54] | -0.01 | [-0.01, -0.00] |
| Non-Hispanic White | 0.21 | [0.14, 0.29] | 0 | [-0.00, 0.00] |
| Female | 0.22 | [0.15, 0.29] | 0 | [-0.00, 0.01] |
| Male | 0.33 | [0.23, 0.42] | 0 | [-0.01, 0.00] |
| Under 18 | 0.21 | [0.08, 0.34] | 0.01 | [-0.00, 0.01] |
| 18 to 25 | 0.34 | [0.21, 0.47] | -0.01 | [-0.02, -0.00] |
| 26 to 35 | 0.41 | [0.31, 0.52] | -0.01 | [-0.01, -0.00] |
| 36 to 45 | 0.32 | [0.22, 0.42] | 0 | [-0.00, 0.01] |
| 46 to 55 | 0.31 | [0.21, 0.41] | 0.01 | [0.00, 0.01] |
| 56 to 65 | 0.19 | [0.09, 0.28] | 0 | [-0.00, 0.01] |
| 66 to 75 | -0.02 | [-0.20, 0.16] | 0.01 | [-0.00, 0.02] |
| 76 or more | 0.04 | [-0.18, 0.26] | 0.01 | [-0.00, 0.02] |
| A month or more of stay-at-home order | 0.34 | [0.25, 0.42] | 0 | [-0.01, 0.00] |
| Less than a month of stay-at-home order | 0.29 | [0.18, 0.41] | 0 | [-0.01, 0.00] |
| Stay-at-home order not<br>implemented | 0.35 | [0.15, 0.54] | -0.01 | [-0.02, 0.00] |

**Table 3.** United States Homicide Absolute Rate Trend Changes Before and After COVID-19 Pandemic Initiation in March 2020, January 2017-December 2022.

|  | Overall Homicide Rate |  | Firearm Homicide Rate |  | Non-Firearm Homicide Rate |  |
| --- | --- | --- | --- | --- | --- | --- |
|  | Absolute<br>immediate<br>change when<br>COVID-19<br>pandemic<br>started | Sustained<br>trend change<br>during<br>COVID-19<br>pandemic | Absolute<br>immediate<br>change when<br>COVID-19<br>pandemic<br>started | Sustained<br>trend change<br>during<br>COVID-19<br>pandemic | Absolute<br>immediate<br>change when<br>COVID-19<br>pandemic<br>started | Sustained<br>trend<br>change<br>during<br>COVID-19<br>pandemic |
| **National** | 1.98<br>[1.49, 2.47] | 0<br>[-0.03, 0.02] | 1.73<br>[1.21, 2.24] | -0.01<br>[-0.03, 0.02] | 0.13<br>[0.04, 0.21] | 0.00<br>[-0.00, 0.01] |
| **Hispanic** | 1.20<br>[0.54, 1.85] | 0.06<br>[0.03, 0.09] | 0.98<br>[0.42, 1.54] | 0.05<br>[0.02, 0.08] | 0.24<br>[0.03, 0.45] | 0.01<br>[0.00, 0.02] |
| **Non-Hispanic**<br>**American**<br>**Native** | 1.27<br>[-1.40, 3.95] | -0.05<br>[-0.18, 0.08] | 1.1<br>[-0.54, 2.73] | -0.06<br>[-0.13, 0.02] | 0.15<br>[-1.53, 1.82] | 0.00<br>[-0.08, 0.08] |
| **Non-Hispanic**<br>**Asian** | 0.12<br>[-0.18, 0.42] | -0.01<br>[-0.02, 0.01] | 0.09<br>[-0.14, 0.32] | -0.01<br>[-0.02, 0.00] | 0.04<br>[-0.17, 0.25] | 0<br>[-0.01, 0.01] |
| **Non-Hispanic**<br>**Black** | 10.09<br>[7.55, 12.63] | -0.12<br>[-0.25, 0.01] | 9.39<br>[6.79, 11.98] | -0.11<br>[-0.25, 0.02] | 0.35<br>[-0.01, 0.71] | 0.00<br>[-0.02, 0.02] |
| **Non-Hispanic**<br>**White** | 0.57<br>[0.36, 0.78] | 0<br>[-0.01, 0.01] | 0.5<br>[0.30, 0.70] | 0<br>[-0.01, 0.01] | 0.04<br>[-0.04, 0.12] | 0<br>[-0.00, 0.00] |
| **Female** | 0.52<br>[0.36, 0.69] | 0<br>[-0.00, 0.01] | 0.52<br>[0.37, 0.68] | 0<br>[-0.00, 0.01] | -0.02<br>[-0.10, 0.06] | 0.00<br>[-0.00, 0.01] |
| **Male** | 3.44<br>[2.56, 4.32] | -0.01<br>[-0.06, 0.03] | 2.97<br>[2.05, 3.89] | -0.02<br>[-0.07, 0.03] | 0.27<br>[0.15, 0.40] | 0.01<br>[-0.00, 0.01] |
| **Under 18** | 0.51<br>[0.22, 0.79] | 0.01<br>[0.00, 0.03] | 0.59<br>[0.35, 0.83] | 0.01<br>[-0.00, 0.02] | -0.09<br>[-0.21, 0.03] | 0.01<br>[0.00, 0.01] |
| **18 to 25** | 5.29<br>[3.40, 7.19] | -0.08<br>[-0.18, 0.02] | 5.2<br>[3.41, 6.98] | -0.08<br>[-0.17, 0.01] | 0.14<br>[-0.07, 0.36] | 0<br>[-0.01, 0.01] |
| **26 to 35** | 5.19<br>[4.02, 6.37] | -0.05<br>[-0.11, 0.00] | 4.78<br>[3.58, 5.98] | -0.05<br>[-0.11, 0.01] | 0.26<br>[0.02, 0.50] | 0<br>[-0.01, 0.01] |
| **36 to 45** | 2.67<br>[1.91, 3.42] | 0.01<br>[-0.02, 0.05] | 2.28<br>[1.58, 2.98] | 0.01<br>[-0.02, 0.05] | 0.36<br>[0.14, 0.59] | 0<br>[-0.01, 0.01] |
| **46 to 55** | 1.51<br>[1.03, 1.99] | 0.02<br>[-0.00, 0.04] | 1.2<br>[0.76, 1.64] | 0.01<br>[-0.01, 0.03] | 0.28<br>[0.05, 0.50] | 0.01<br>[-0.00, 0.02] |
| **56 to 65** | 0.64<br>[0.32, 0.95] | 0<br>[-0.01, 0.02] | 0.44<br>[0.22, 0.66] | 0.01<br>[-0.00, 0.02] | 0.2<br>[-0.00, 0.41] | 0<br>[-0.01, 0.01] |
| **66 to 75** | -0.05<br>[-0.39, 0.30] | 0.01<br>[-0.00, 0.03] | 0.02<br>[-0.20, 0.24] | 0.01<br>[0.00, 0.02] | -0.07<br>[-0.32, 0.18] | 0<br>[-0.01, 0.01] |
| **76 to older** | -0.02<br>[-0.39, 0.35] | 0.01<br>[-0.01, 0.03] | 0.11<br>[-0.09, 0.31] | -0.01<br>[-0.02, 0.00] | -0.13<br>[-0.41, 0.14] | 0.02<br>[0.00, 0.03] |
| **Month or More** | 1.95<br>[1.52, 2.38] | 0<br>[-0.02, 0.02] | 1.63<br>[1.15, 2.12] | -0.01<br>[-0.03, 0.02] | 0.14<br>[0.05, 0.23] | 0<br>[0.00, 0.01] |
| **Less than a**<br>**Month** | 2.36<br>[1.55, 3.18] | -0.02<br>[-0.06, 0.02] | 2.35<br>[1.59, 3.10] | -0.02<br>[-0.06, 0.02] | 0.01<br>[-0.19, 0.20] | 0<br>[-0.01, 0.01] |
| **Not**<br>**Implemented** | 1.42<br>[0.70, 2.14] | -0.04<br>[-0.07, -0.00] | 1.19<br>[0.59, 1.79] | -0.03<br>[-0.06, -0.00] | 0.22<br>[-0.13, 0.56] | -0.01<br>[-0.02, 0.01] |

Prior to the pandemic, most of these groups had stable, constant homicide mortality rate trends; however, during the pandemic every group experienced an immediate increase in homicide mortality rates (Figures 1-4; Table 2). The sustained trend changes during the pandemic compared to pre-pandemic for most groups were minimal or slightly declining (Figures 2-4). For all groups examined, increases in homicide mortality rates coincided with increases in firearm-related homicide mortality rates, indicating that firearms were the primary method by which homicide mortality rates increased.

**Figure 2.**
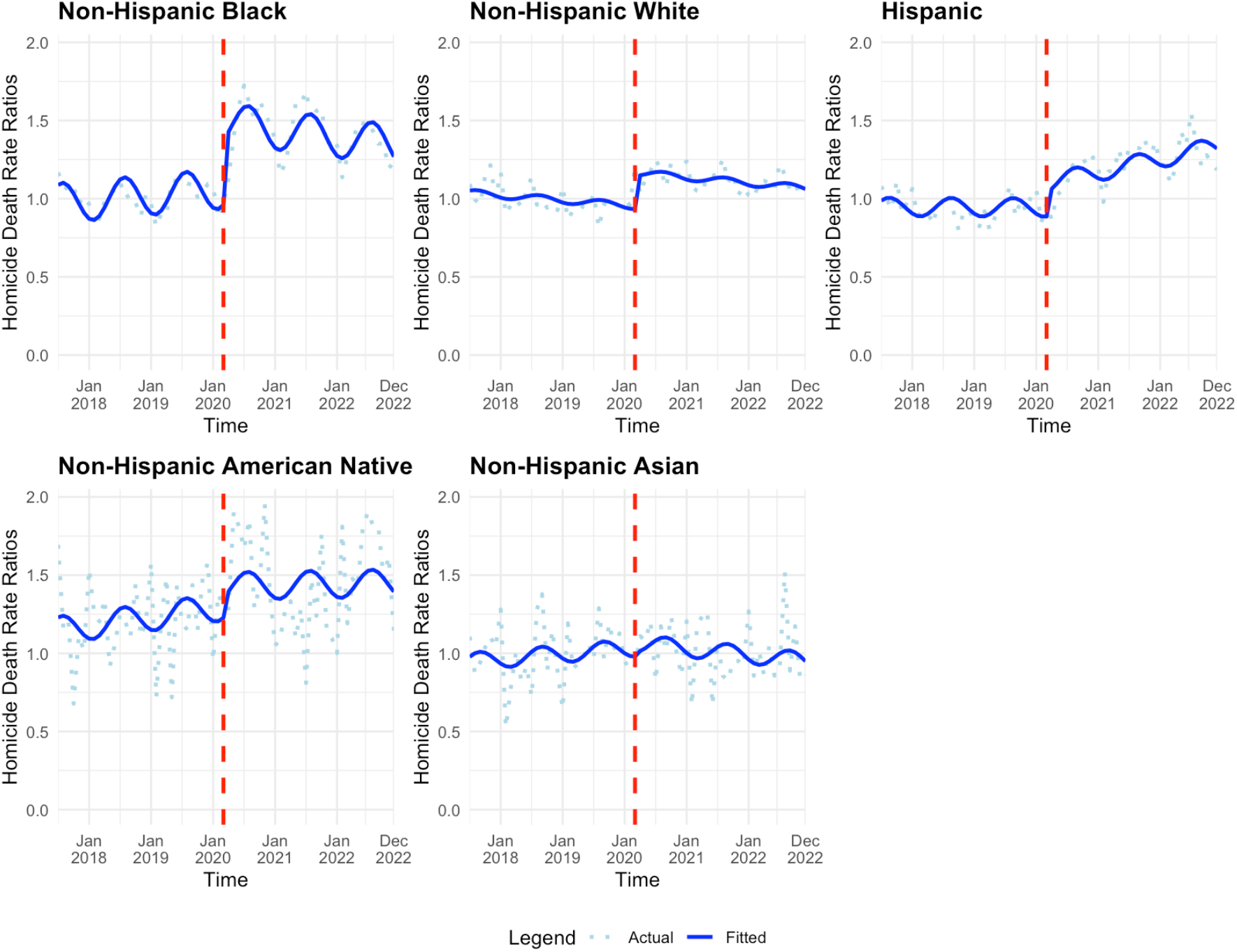
National Homicide Deaths by Race and Ethnicity (a) Rates and (B) Rate Ratios Over Time per 100,000 PY, July 2017-December 2022

Restricting by race, we observe that the NH Black population followed by the NH White population had the greatest relative increases to their homicide mortality rates immediately after the COVID-19 pandemic onset compared to their pre-pandemic homicide mortality rates (Table 2). The NH Black population’s rates increased by 42% (95% CI: 30%, 54%), and the NH White population’s rates increased by 21% (95% CI: 14%, 29%).

Comparing males to females, males the had a higher immediate change to their homicide mortality rate when the pandemic started (Figure 3; Table 2). Male homicide mortality rates increased by 33% (95% CI: 23%, 42%). On the other hand, female homicide mortality rates increased by 22% (95% CI: 15%, 29%).

**Figure 3.**
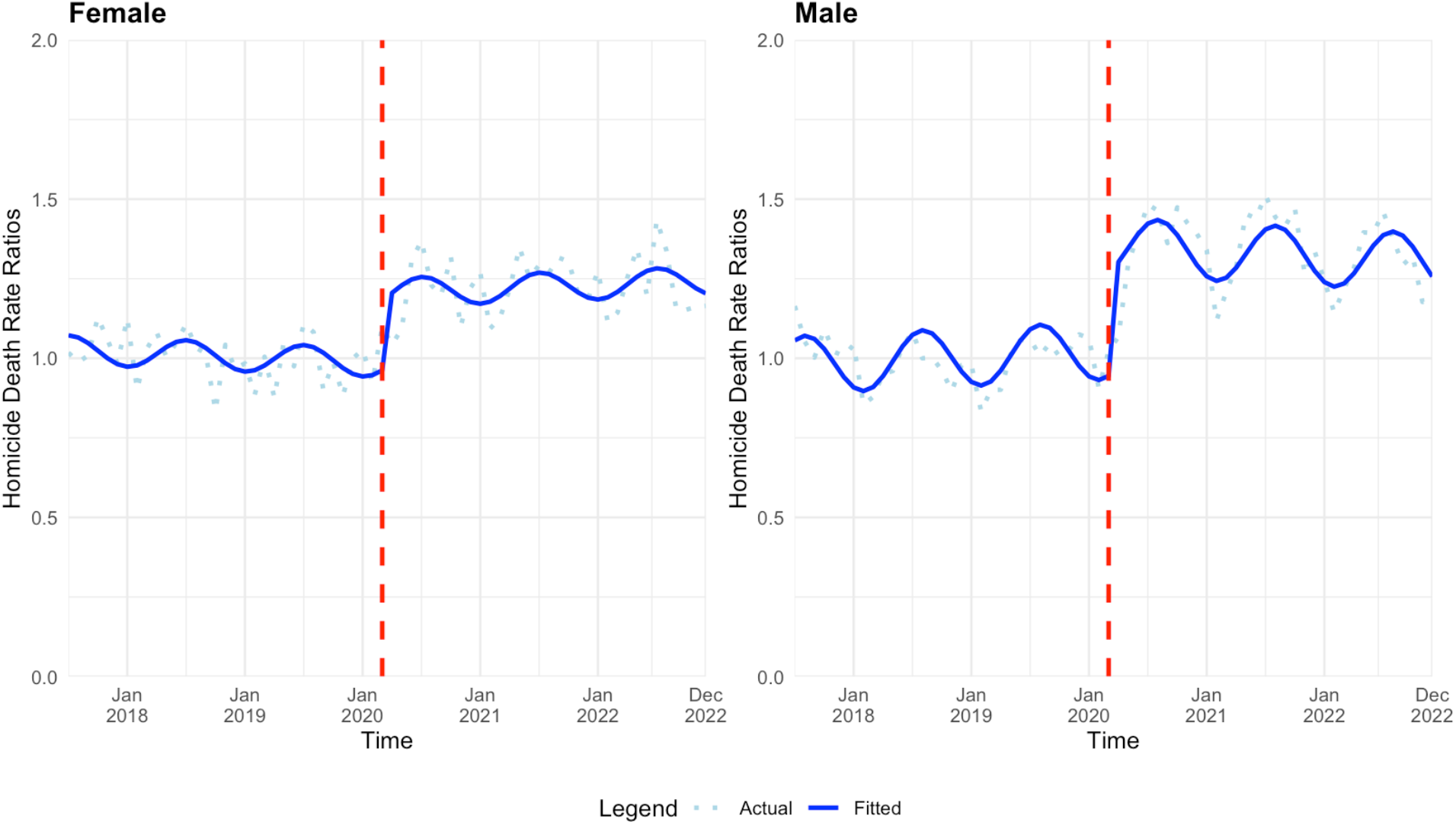
National Homicide Deaths by Sex (a) Rates and (B) Rate Ratios Over Time per 100,000 PY, July 2017-December 2022

In terms of age, individuals 65 and under faced immediate increases in their homicide mortality rates (Figure 4; Table 2). Those in the age group 26-35 years old had the greatest immediate increase to their homicide mortality rates, 41% (95% CI: 31%, 52%) followed by 18-to 25-year-olds (34% (95% CI: 21%, 47%)) then 36 to 45 (32%, 95% CI: 22%, 42%) and 46-to 55-year-olds (31%, 95% CI: 21%, 41%). Those in age groups 66 and older faced little or no change to their homicide mortality rates.

**Figure 4.**
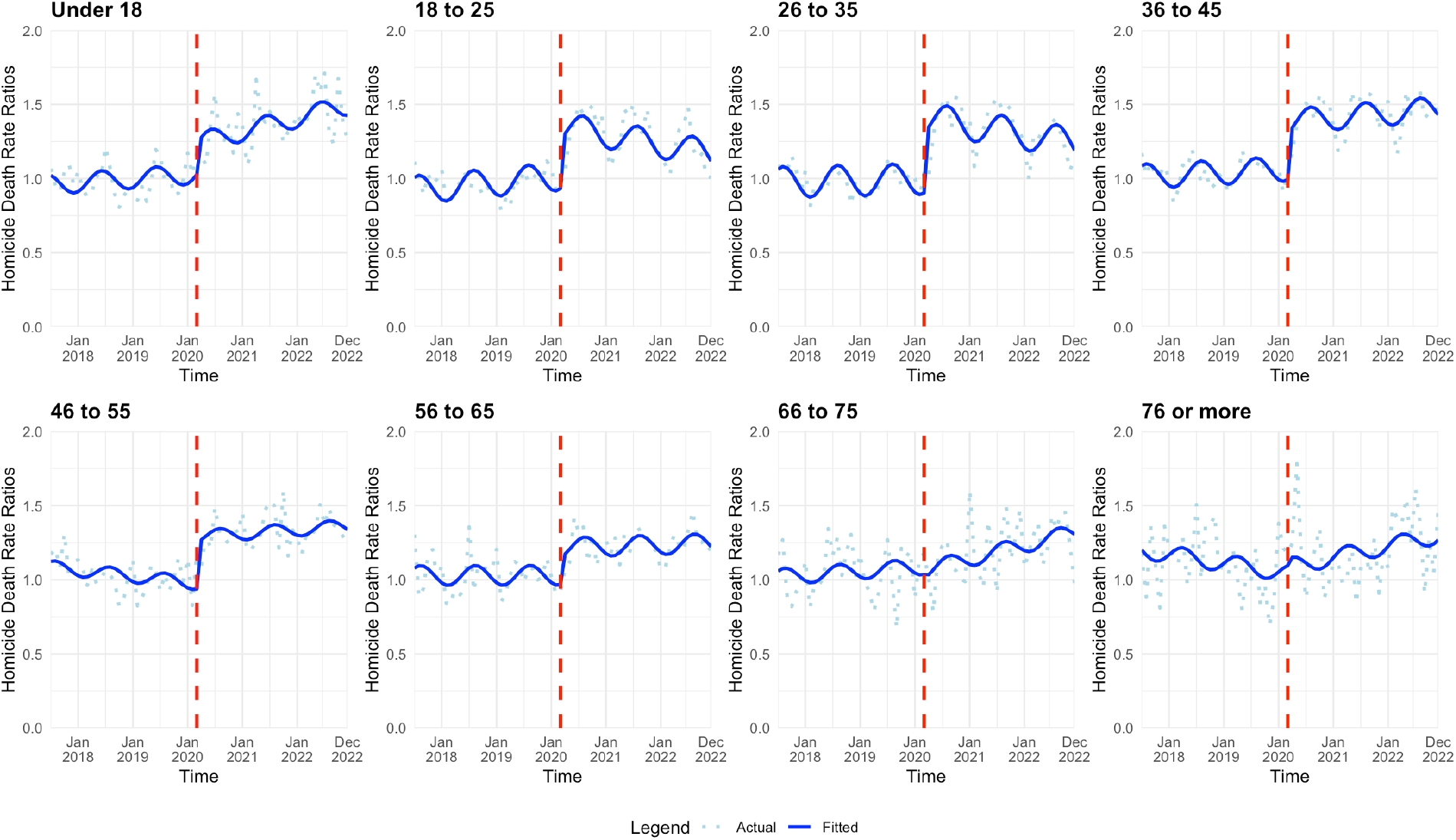
National Homicide Deaths by Age Group (a) Rates and (B) Rate Ratios Over Time per 100,000 PY, July 2017-December 2022

All three SAHO groups experienced similar increases to their homicide mortality rates (Figure 5; Table 2). States with a SAHO not implemented had the greatest increase in mortality rate (35% (95% CI: 15%, 54%)), closely followed by states with a month or more of SAHO (34% (95% CI: 25%, 42%), and finally states with SAHO less than a month experienced the lowest increase in homicide mortality rate ratios, increasing by 29% (95% CI: 18%, 41%).

**Figure 5.**
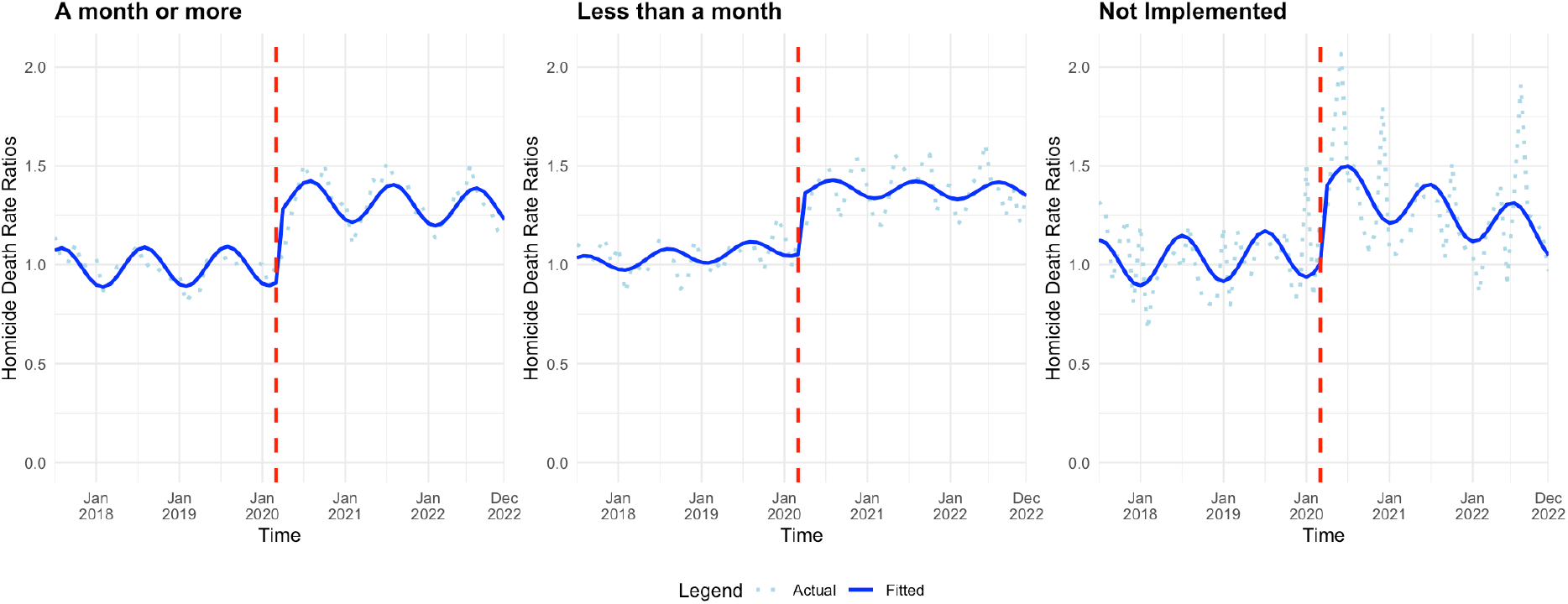
National Homicide Deaths by State Stay at Home Order Length (a) Rates and (B) Rate Ratios Over Time per 100,000 PY, July 2017-December 2022

## Discussion

Homicide rates immediately increased following COVID-19 pandemic onset and remained relatively stable after their initial increase in the two years following the COVID-19 pandemic, indicating a level change. Prior to pandemic onset, the US homicide mortality rate was stable. However, immediately after the COVID-19 pandemic began, the overall homicide mortality rate ratio increased by 32% (95% CI: 23%, 41%), which persisted through December 31, 2022. Most stratified analyses revealed similar increases to homicide rates immediately after the pandemic. Non-Hispanic Black Americans followed by NH White Americans experienced the some of the greatest immediate increases to their homicide mortality ratio rates, respectively. All age groups under 65 experienced immediate increases in their homicide mortality rates. Firearm-related homicides increased also increased to a greater extent than non-firearm-related homicides. Similar relative increases were observed in states regardless of SAHO. Like overall trends, each of these immediate changes to rate ratios persisted until the end of 2022 without additional trend changes.

There are some potential explanations for why we observe this increase in homicide mortality immediately after the COVID-19 pandemic. Prior research has demonstrated that homicide rates are sensitive to large-scale crises such as natural disasters and pandemics.^19,20^ These crises induce states of stress and exacerbate existing health disparities that put vulnerable people at amplified risk. For example, hurricanes intensify depression, post-traumatic stress, substance use, and anxiety as well as economic challenges and deteriorated social order, all of which were observed during the COVID-19 pandemic.^4,21^ Research has also linked hurricanes with increased rates of violent injury and suicide, suggesting that COVID-19, as a state of emergency, may have had a similar impact.^19,22^ Some evidence of this was observed at the start of the pandemic, with an immediate increase in firearm sales, emergency department visits from firearm-related violence, and homicide deaths in the United States.^23–25^ Early reports also noted an increase in calls to 911 and domestic violence hotlines regarding intimate partner violence. ^20,21^ A later study found a 30% increase in homicide death rate between 2019 (4.6 deaths per 100,000 population) to 2020 (6.1 per 100,000 population), with homicide rates being largely driven by firearm related homicides.^11,25^ More recent studies have found firearms sales increased between March 2020 through 2021 due to social and political unrest which may have contributed to the observed increases in firearm homicide rates following COVID-19 onset.^8^ Additionally, while prior literature suggests that lifestyle changes due to SAHOs may have had impacts on violent crime,^28–30^ including homicide, at the city level,^31^ relative homicide rates were not observed to have differed significantly between states with varying durations of SAHOs. This may be due to heterogeneity in the terms of the orders, differences in state-level urbanicity, and the fact that most states implemented orders lasting more than 30 days. Moreover, other studies found that intimate partner violence rates also increased while rates of crimes like robbery and larceny declined, indicating that SAHOs may have differentially impacted different types of crime.

Black Americans faced some of the greatest burdens during the COVID-19 pandemic, as pre-existing systemic racism and disparities were intensified. NH Black Americans had the greatest homicide rates both before and after the start of the pandemic. However, compared to all other racial and ethnic groups, NH Black Americans experienced the greatest relative increase in homicide rates, which remained elevated through 2022. ^29^ This larger relative increase in homicide rate had significant impacts on the Black population with homicide being a leading contributing factor for decreases in life expectancy for Black Americans during the pandemic period.^33,34^ Reasons for this may include that this group was disproportionately infected and died from COVID-19 and also faced greater socio-economic pressures during the pandemic.^34^ Additionally, Black Americans also faced growing anti-Black bias through increased discussions and protests due to the widely shared police brutality incidents against Black Americans, resulting in further social unrest and stress in communities. These factors likely increased interpersonal stress and tension, contributing to higher levels of violence. This pattern is reflected in research examining gun violence following the murder of George Floyd in Minneapolis found increased rates of gun violence in areas closer to protests, particularly historically Black communities.^35,36^ Overall, the additional burdens this population faced during the pandemic may have also contributed to their relatively large increase in homicide rates compared to other racial and ethnic populations. The unique characteristics of the relative increase of NH Black American homicide rates during and following the pandemic should be explored further in future research.

This study is not without limitations. First, this study only considers homicides, which is an extreme and rare type of violence. Other forms of fatal and non-fatal violence that were not assessed. Second, categorizations used in demographic racial and ethnic sub-analyses may not be accurately captured in the data, especially for the Hispanic population, and analysis for multi-racial populations were not possible. Third, while COVID may have served as an impetus for the social disruption and tension that led to increased homicide rates, certain events such as the nationwide Black Lives Matter protests, 2020 US election, and January 6 United States Capitol attack may be more closely linked to immediate and sustained increases to homicide rates.

By using quasi-experimental approach to observe the impact of the pandemic’s onset on homicide rates, we observe that homicide rates immediately increased following the COVID-19 pandemic and remained relatively stable in the two years following. We also found that firearm homicides in particular increased during this period. Public health practitioners can use this information to advocate for violence prevention efforts to occur in tandem with other public health offers during states of emergency, including future pandemics, hurricanes, or other large-scale crises, with a particular focus on gun-specific prevention strategies. Moreover, these findings indicate that pandemics and potentially other emergent situations may have sustained, but time-limited, effect on violence, given that homicide rates appeared to have decreased in 2023. ^35^ These higher rates of homicide mortality rates relative to pre-pandemic levels should be of concern and be considered a significant cascading effect of the pandemic that must be addressed. Future research should examine the trend changes to the characteristics of homicides, such as the circumstances in which they occurred, the characteristics of offenders, and regional variations in impact, and other forms of violence that occurred during the COVID-19 pandemic.

## Data Availability

All data used in this study were obtained through a request for Restricted-Use National Vital Statistics System data.

