## Supplemental Materials for "COVID-19 onset, stay-at-home orders, and racialized inequities in homicide mortality across the US"

### State lockdown orders^14^

| **State** | **Lockdown Order Dates** | **Number of Days in Lockdown** | **Lockdown Group** | **Official name of order** |
| --- | --- | --- | --- | --- |
| Alabama | April 4- April 30 | 26 | Less than a month | Suspend certain public gatherings |
| Alaska | March 28 - April 24 | 27 | Less than a month | Shelter-in-place |
| Arizona | March 31 - May 15 | 45 | A month or more | Stay home, stay healthy, stay connected |
| Arkansas | None | 0 | Not Implemented | N/A |
| California | March 19 - August 28 | 162 | A month or more | Shelter-in-place |
| Colorado | March 26 - April 26 | 31 | A month or more | Stay-at-home |
| Connecticut | March 23 - May 20 | 58 | A month or more | Stay Safe, Stay Home |
| Delaware | March 24 - May 31 | 68 | A month or more | Stay-at-Home |
| Florida | April 2 - May 4 | 32 | A month or more | Stay-at-home |
| Georgia | April 3 - April 30 | 27 | Less than a month | Shelter-in-place |
| Hawaii | March 25 - May 31 | 67 | A month or more | Stay-at-home |
| Idaho | March 25 - April 30 | 36 | A month or more | Stay home |
| Illinois | March 21 - May 29 | 69 | A month or more | Stay-at-Home |
| Indiana | March 24 - May 1 | 38 | A month or more | Stay-at-home |
| Iowa | None | 0 | Not Implemented | N/A |
| Kansas | March 30 - May 3 | 34 | A month or more | Stay home |
| Kentucky | March 26 - June 29 | 95 | A month or more | Stay healthy at home |
| Louisiana | March 23 - May 15 | 53 | A month or more | Stay-at-Home |
| Maine | April 2 - May 31 | 59 | A month or more | Stay-at-home |
| Maryland | March 30 - May 15 | 46 | A month or more | Stay-at-home |
| Massachusetts | March 24 - May 18 | 55 | A month or more | Stay-at-home |
| Michigan | March 24 - June 1 | 69 | A month or more | Stay Home, Stay Safe |
| Minnesota | March 27 - May 17 | 51 | A month or more | Stay-at-home |
| Mississippi | April 3 - April 27 | 24 | Less than a month | Shelter-in-place |
| Missouri | April 6 - May 3 | 27 | Less than a month | Stay Home Missouri |
| Montana | March 28 - April 26 | 29 | Less than a month | Stay-at-home |
| Nebraska | None | 0 | Not Implemented | N/A |
| Nevada | April 1 - May 15 | 44 | A month or more | Stay-at-home |
| New Hampshire | March 27 - June 15 | 80 | A month or more | Stay-at-home 2.0 |
| New Jersey | March 21 - June 9 | 80 | A month or more | Stay-at-home |
| New Mexico | March 24 - November 30 | 251 | A month or more | Stay-at-home |
| New York | March 20 - June 27 | 99 | A month or more | New York State on PAUSE |
| North Carolina | March 30 - May 22 | 53 | A month or more | Stay-at-home |
| North Dakota | None | 0 | Not Implemented | N/A |
| Ohio | March 23 - May 19 | 57 | A month or more | Stay-at-home |
| Oklahoma | April 1 - May 6 | 35 | A month or more | Safer at home |
| Oregon | March 23 - June 19 | 88 | A month or more | Stay-at-home |
| Pennsylvania | April 1 - June 4 | 64 | A month or more | Stay-at-home |
| Rhode Island | March 28 - May 8 | 41 | A month or more | Stay-at-home |
| South Carolina | April 7 - May 4 | 27 | Less than a month | Home or work |
| South Dakota | None | 0 | Not Implemented | N/A |
| Tennessee | March 31 - April 30 | 30 | Less than a month | Safer at home |
| Texas | April 2 - April 30 | 28 | Less than a month | Statewide essential services and activities protocols |
| Utah | None | 0 | Not Implemented | N/A |
| Vermont | March 24 - May 15 | 52 | A month or more | Stay-at-home |
| Virginia | March 30 - May 29 | 60 | A month or more | Stay-at-home |
| Washington | March 24 - May 31 | 68 | A month or more | Stay-at-home |
| West Virginia | March 24 - May 4 | 41 | A month or more | Stay-at-home |
| Wisconsin | March 25 - May 13 | 49 | A month or more | Safer at Home |
| Wyoming | None | 0 | Not Implemented | N/A |

### Absolute Scale Analysis by Strata

The greatest absolute immediate change following the start of the pandemic among the NH Black population with an increase of 10.09 homicide deaths/100,000 PY (95% CI: 7.55, 12.63), going from 22.93 homicide deaths/100,000 PY pre-pandemic to 32.44 homicide deaths/100,000 PY post-pandemic. Like the national model, this trend persisted until the end of 2022 with a sustained decline trend change of 0.12 homicide deaths/100,000 PY (95% CI: -0.25, 0.01) with seasonal variations (Figure 1).

The next highest absolute immediate change after the pandemic began was among Native American people. Their homicide rate pre-pandemic was lower at 10.81 homicide deaths/100,000 PY. With the onset of the pandemic, there was an increase of 1.27 homicide deaths/100,000 PY (95% CI: -1.40, 3.95). This trend declines by 0.05 homicide deaths/100,000 PY 95% CI: -0.18, 0.08) among Native American/Indigenous people from the pandemic onset to the end of the study period.

Hispanic people had the next highest absolute immediate change to their homicide rates (1.20 deaths per 100,000 PY (95% CI: 0.54, 1.85)). However, the Hispanic population was the only group to have a sustained increasing homicide rate trend from March 2020 to the end of the study period with an increase of 0.06 homicide deaths/100,00 PY (95% CI: 0.03, 0.09).

We observed that while the homicide rate among NH Asian Americans did not increase as dramatically as the aforementioned racial and ethnic groups following the pandemic onset in March 2020 (an increase of 0.12 homicide deaths/100,000 PY (95% CI: -0.18, 0.42).

Comparing males to females, males the highest absolute immediate change to their homicide rates when the pandemic started, with an increase of 3.44 homicide deaths/100,000 PY (95% CI: 2.56, 4.32). Females had an increase of 0.52 additional homicide deaths/100,000 PY (0.36, 0.69). Males had a sustained decreasing trend of 0.01 fewer homicide deaths/100,000 PY (95 CI: -0.06, 0.03) while females had a minimally increasing trend (0 additional homicide deaths/100,000 PY (95 CI: 0.00, 0.01)) (Figure 2).

Individuals aged 18 to 25 years, followed by those aged 26 to 35 years, experienced a larger increase in homicide mortality than any other age groups (Figure 3). Pre-pandemic, 18- to 25-year-olds had the highest homicide mortality rate at 13.96/100,000 PY with 26- to 35-year-olds following at 11.86/100,000 PY. With pandemic onset, 18- to 25-year-old homicide mortality rate increased by 5.29/100,000 PY (95% CI: 3.40, 7.19) to 18.40 homicide deaths per 100,000 PY and 26- to 35-year-old homicide mortality rate increased by 5.19/100,000 PY (95% CI: 4.02, 6.37) to 15.99. However, in the months following COVID-19 onset, the sustained trend had a decrease of -0.08 homicide deaths/100,000 PY (95% CI: -0.18, 0.02) and -0.05/100,000 PY (95% CI: -0.11, 0.00) among 18- to 25-year-olds and 26- to 35-year-olds respectively. This trend persisted until the end of 2022 with seasonal variations.

In January 2017, prior to the COVID-19 pandemic, states with less than a month of lockdown had the highest homicide mortality rate at 7.99/100,000 PY and states that did not implement a lockdown had the lowest homicide rate at 4.11/100,000 PY. COVID-19-related immediate homicide mortality increases occurred in all states regardless of SAHO with states with less than a month of lockdown having the largest trend increase of 2.36/100,000 PY (95% CI: 1.55, 3.18). All states, trends persisted until the end of 2022 with mainly minimal trend changes but with seasonal changes (Figure 5).

#### Figure 1. National Homicide Deaths by Race and Ethnicity (a) Rates and (B) Rate Ratios Over Time per 100,000 PY, July 2017-December 2022

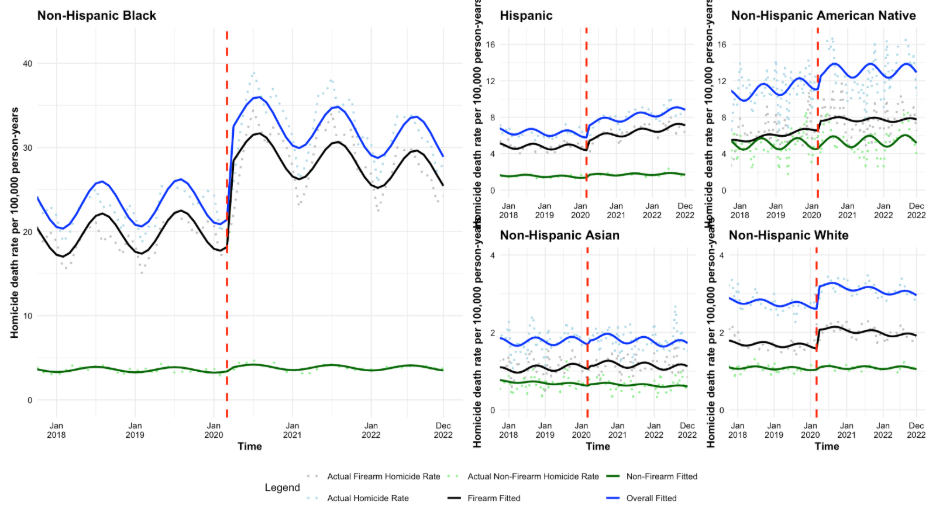

*Figure 2. National Homicide Deaths by Sex (a) Rates and (B) Rate Ratios Over Time per 100,000 PY, July 2017-December 2022*

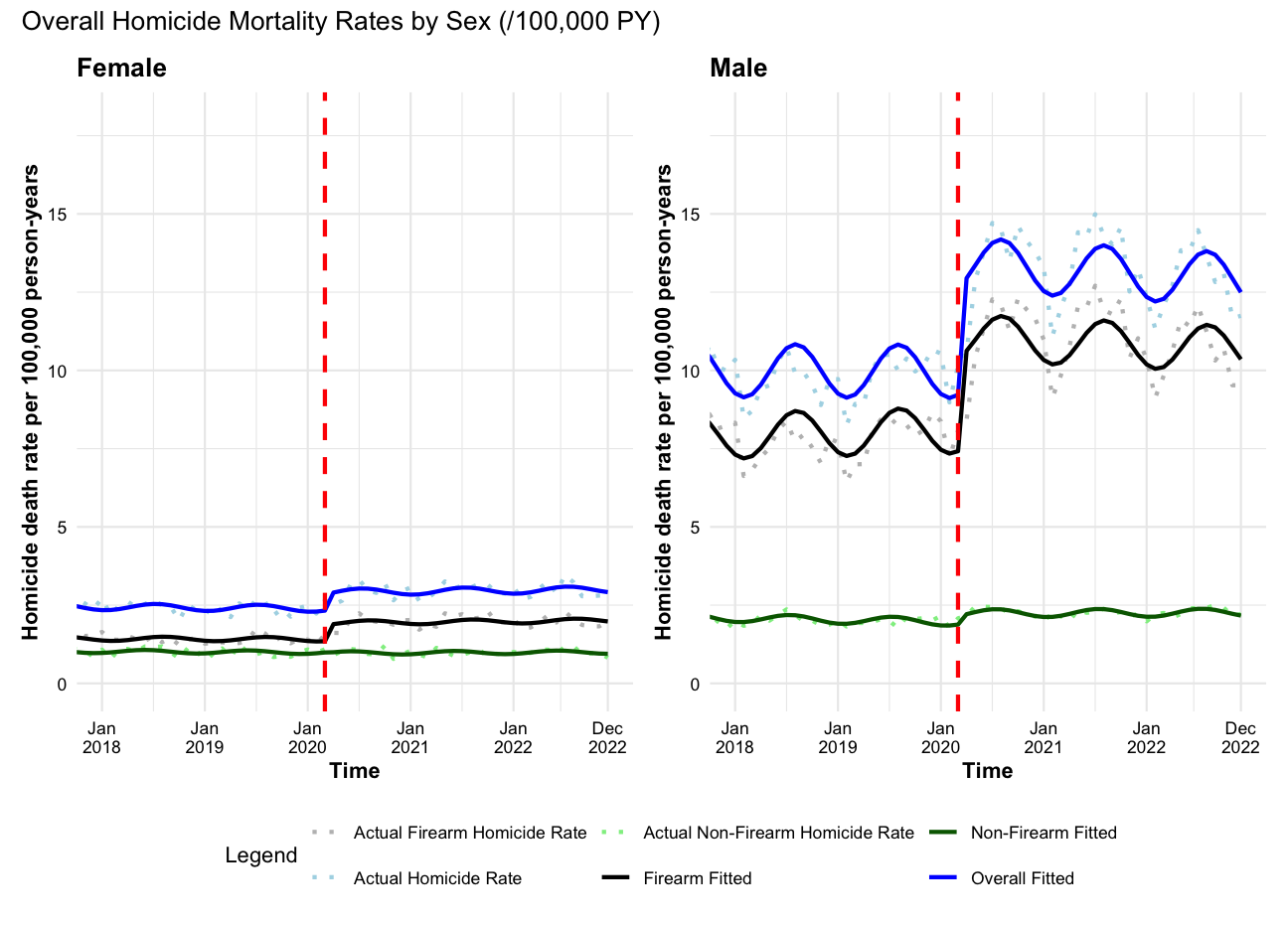

#### Figure 3. National Homicide Deaths by Age Group (18-45) (a) Rates and (B) Rate Ratios Over Time per 100,000 PY, July 2017-December 2022

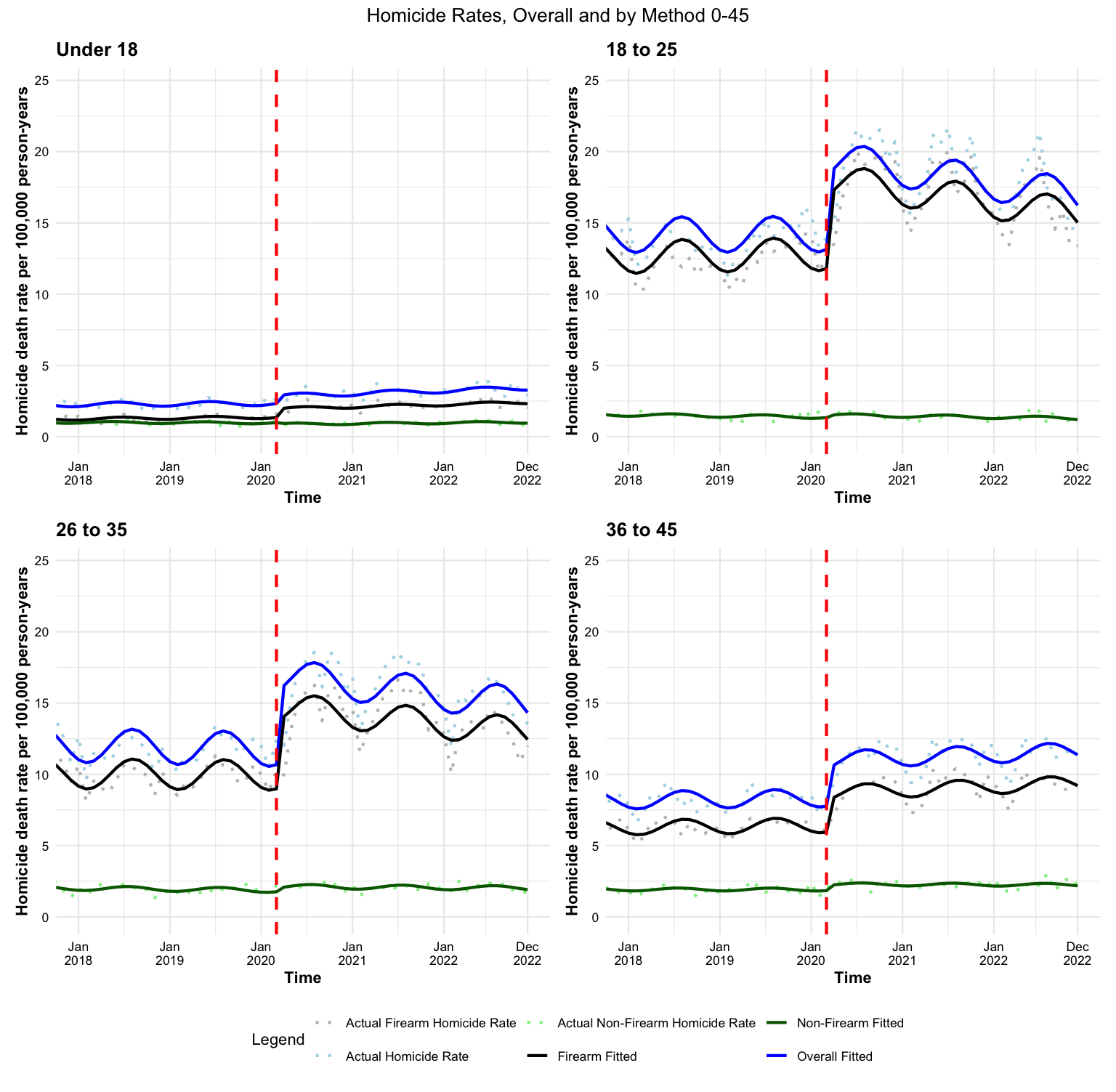

*Figure 4. National Homicide Deaths by Age Group (46+) (a) Rates and (B) Rate Ratios Over Time per 100,000 PY, July 2017-December 2022*

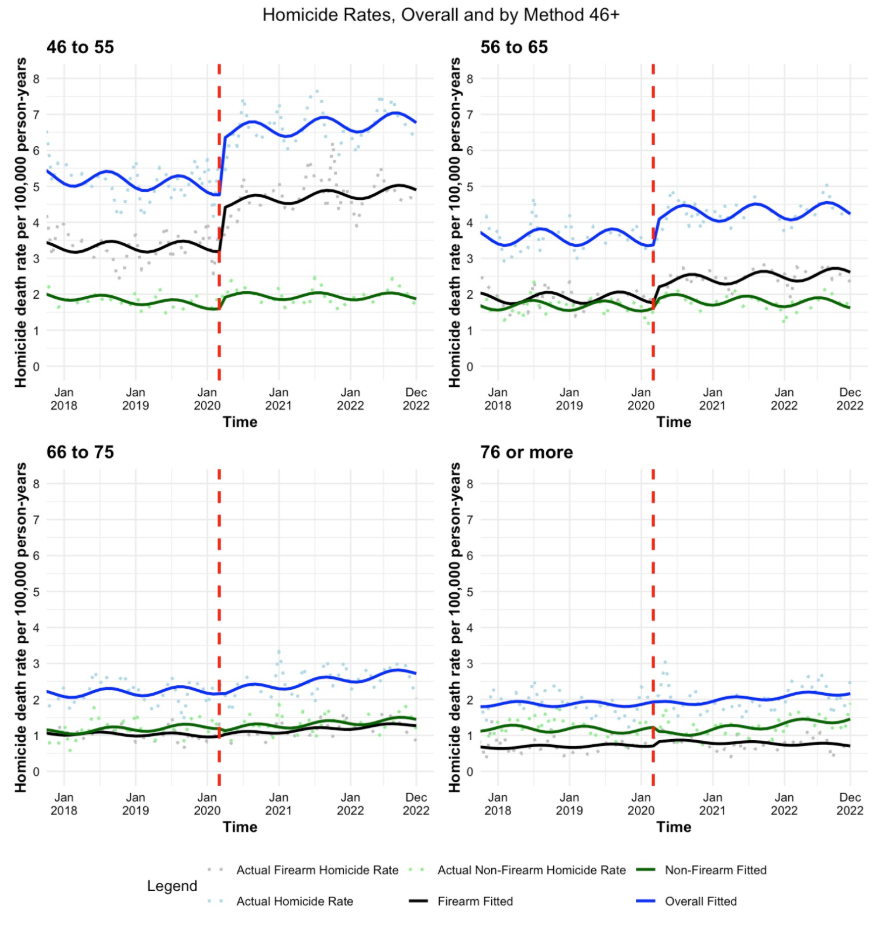

#### Figure 5. National Homicide Deaths by State Stay at Home Order Length (a) Rates and (B) Rate Ratios Over Time per 100,000 PY, July 2017-December 2022

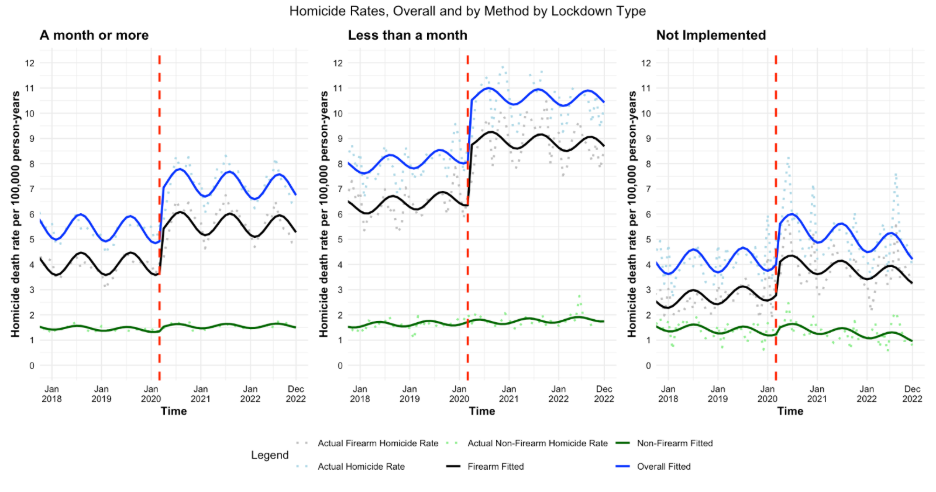
